# Evaluation of an ultra-rapid antibiotic susceptibility testing method on positive blood cultures with *Escherichia coli*

**DOI:** 10.1101/2021.12.14.21267046

**Authors:** Özden Baltekin, Alexander T. A. Johnsson, Alicia Y. W. Wong, Kajsa Nilsson, Berivan Mert, Lovisa Söderberg, Erik Wistrand-Yuen, Volkan Özenci

## Abstract

Blood stream infection (BSI) is related to high mortality and morbidity. Early antimicrobial therapy is crucial in treating patients with BSI. The most common Gram-negative bacteria causing BSI is *Escherichia coli*. Targeted effective treatment of patients with BSI is only possible if it is based on antibiotic susceptibility testing (AST) data after blood culture positivity. However, there are very few methods available for rapid phenotypic AST and the fastest method takes 4 h. Here we analyzed the performance of a 30 min ultra-rapid method for AST of *E. coli* directly from positive blood cultures (BC). In total, 51 positive BC with *E. coli* were studied, and we evaluated the ultra-rapid method directly on positive BC as well as on *E. coli* colonies cultured on agar plates. The results obtained by the new method were compared with disk diffusion.

The method provided accurate AST result in 30 min to Ciprofloxacin and Gentamicin for 92% and 84% of the positive BC samples, respectively. For *E. coli* isolates retrieved from agar plates, 86% and 96% of the AST results were accurate for Ciprofloxacin and Gentamicin, respectively, after 30 min of assay time. When time to result was modulated in-silico from 30 to 60 minutes for the agar plate samples, accuracy of AST results went up to 92% for Ciprofloxacin and to 100% for Gentamicin.

The present study shows that the method is reliable and delivers ultra-rapid AST data in 30 minutes directly from positive BC and as well as from agar plates.

## INTRODUCTION

Bloodstream infections (BSI) are associated with high morbidity, mortality, and high health-care related costs (Angus et al., 2003). Early identification and susceptibility testing of the causative agent have shown that improvement in the disease outcomes [1, 2]. However, conventional identification and susceptibility testing methods can take 48-72 h to be completed [3].

Recent improvements in rapid identification methods have enabled identification of microorganisms to be performed directly from positive blood cultures following a short 4h incubation on solid media [4]. In contrast, there are limited possibilities for rapid antimicrobial susceptibility testing (AST). Molecular AST methods, including FilmArray (bioMérieux, Inc, Hazelwood, Mo) and Verigene (Nanosphere, Northbrook, IL), can perform direct AST from positive blood culture (BC) bottles and deliver results in 60-120 minutes. However, these molecular methods can only perform AST for a limited number of antimicrobials. Other rapid phenotypic AST methods come with drawbacks. The VITEK 2 (bioMeriéux) and Phoenix 100 (BD Biosciences) systems require more hand-on time as the setup involves dilution of pure microbe colonies to a specified McFarland standard [5]. The Accelerate Pheno™ system improves on this by automating both identification and AST directly from positive BCs, but suffers from issues stemming from problems with identification of the microorganism that affect the downstream AST result [6]. Other automated AST rapid methods circumvent this issue by relying on microbe identity to be provided separately, such as the ASTar® (Q-linea) [7] and dRAST™ (Quantamatrix) [8]. However, these rapid phenotypic AST methods provide results in 6-8 h. As this is close to the end of the laboratory- and clinical-working shift, any possible escalation and de-escalation of the patient’s antibiotic treatment might be performed the day after as in the case of conventional AST.

It is important to establish AST methods with very short turn-around time after the BC bottles signal positive so that the necessary adjustment of empirical antimicrobial treatment can be done the same day. Recently we developed a 30 min ultra-rapid phenotypic AST method based on rapid isolation and real time single cell phenotyping of bacteria growth using a single microfluidic device. The ultra-rapid phenotypic AST method has been shown to have high performance in providing ultra-rapid AST results when tested on clinical isolates of uropathogenic *Escherichia coli* spiked into urine substitute (Baltekin et al., 2017). The present study aims to analyze the performance of the ultra-rapid method for AST of BSI-causing *E. coli* on both positive BCs and on *E. coli* colony isolates.

## MATERIALS AND METHODS

### Study design

The present pilot study focuses on rapid AST on positive blood cultures with *E. coli* and was performed in two parts. First part is dedicated to evaluation of the ultra-rapid AST with positive blood culture samples with a 30 min turnaround time. The second part aimed at evaluating the method with colonies retrieved from agar plates with 30 min turnaround time as well as analyzing the effect of longer turnaround times from 30 to 60 minutes.

### Sample Collection

Prospective clinical BC samples collected by standard protocols at Karolinska University Hospital Huddinge by standard protocols in BacT/Alert-FA Plus and BacT/Alert-FN Plus BC bottles (bioMérieux, Marcy-l’Étoile, France) were used for the study. All BC bottles were incubated at the Clinical Microbiology Laboratory in Karolinska University Hospital (Huddinge, Sweden) in the automated BacT/ALERT 3D blood culture system (bioMérieux, Marcy-l’Étoile, France) until positivity, or for a maximum of 5 days. Gram staining was performed on BC bottles that signaled positive. Identification of the bacteria in positive BCs were done using short-term culture followed by MALDI-TOF MS [4]. Only BCs with *E. coli* were used for this study. In addition, the *E. coli*-positive blood cultures were also subcultured onto blood agar plates (The Substrate Unit, Karolinska University Hospital, Huddinge, Sweden) and the *E. coli* isolates were stored at –80 °C in glycerol freezing medium (The Substrate Unit, Karolinska University Hospital, Huddinge, Sweden).

### Description of ultra-rapid AST system

The ultra-rapid AST system was developed for investigational use and provided by Astrego Diagnostics AB (Uppsala, Sweden), and compromises of disposable AST cartridges and accompanying analyzer as described previously [9]. The AST cartridge contains a liquid sample reservoir, liquid media reservoirs and a micro-nanofluidic chip (version CD4-Glass), and one AST cartridge was used for each sample of the study. The analyzer prototype was made in collaboration between 77 Elektronika Ltd. (Budapest, Hungary) and Astrego Diagnostics AB (Uppsala, Sweden), to combine functionalities for microscopy and pressure driven flow control into a small tabletop unit [9]. In brief, the analyzer has a slot used for the insertion of a sample-loaded cartridge and is controlled from a software user interface on a connected laptop computer to perform the test. The analyzer maintained the temperature of the inserted cartridge at 36±1 °C, performed all the assay sequences by pressure driven flow control of liquids through the cartridge, and acquired data through time-lapse phase contrast microscopy imaging. The system was calibrated to susceptibility breakpoint using *E. coli* ATCC 25922, and *E. coli* isolates provided by Astrego Diagnostics AB with known minimum inhibitory concentration (MIC) values for Gentamicin and Ciprofloxacin (Supp. Table 1).

### Sample processing for the ultra-rapid AST system

The ultra-rapid AST system was tested directly on *E. coli*-positive blood cultures as well as the frozen clinical *E. coli* isolates. Sample processing from positive BC bottles involved aspirating 3 ml of culture fluid, centrifugation of the culture fluid for 5 min at 50 RCF to sediment any large particles of resin beads and blood clots that would be present in the BC bottle. 500 μl of the top culture fluid after centrifugation was loaded into the sample reservoir of the cartridge using a micropipette, thus avoiding the sedimented particulate matter. The frozen *E. coli* isolates were prepared by retrieving them from the freezer and plating them out on blood agar plates (The Substrate Unit, Karolinska University Hospital, Huddinge, Sweden). The resulting pure colony growth was used in the present study. Ten colonies from each isolate were suspended in 11 ml MHB2. No density adjustments were performed, and 500 ml of the bacterial suspension was loaded into the sample port of the cartridge using a micropipette.

### Preparation of the AST cartridge

The AST cartridge was prepared by first loading it with pre-diluted aliquots of antibiotics. The following reagents were prepared beforehand, aliquoted to 0.5 ml tubes and stored at −20 °C: Cation Adjusted Mueller Hinton Broth (MHB2) (Sigma-Aldrich, Saint Louis, Missouri, USA), MHB2+Gentamicin (4 ug/mL) (Sigma-Aldrich, Saint Louis, Missouri, USA), MHB2+Ciprofloxacin (0.5 ug/mL) (Sigma-Aldrich, Saint Louis, Missouri, USA), and Surfactant (pluronic F-108542342; Sigma-Aldrich, Saint Louis, Missouri, USA, 0.085% (w/v) final concentration). Reagents were thawed at 37 °C for 5 min, and then vortexed till homogenous just before the assay. The AST cartridge was then loaded with 330 μl of each reagent using a micropipette into their respective liquid media reservoirs in the cartridge.

When the cartridge had been loaded with all reagents and sample, it was inserted into the analyzer for automated AST analysis for 20 min to 60 min. Data analysis method is described in the supplementary material.

### Comparison with reference AST testing

The reference method used is the EUCAST standardized disk diffusion method [10, 11] that is performed in the clinical routine at the Clinical Microbiology Laboratory in Karolinska University Hospital (Huddinge, Sweden). AST results obtained by the reference method for all clinical BC isolates used in the present study were retrieved from the clinic Laboratory Information System (LIS), and results obtained from the ultra-rapid AST system were compared with reference method. Antimicrobial susceptibility for all isolates included in this study was tested by disk diffusion following EUCAST guidelines and breakpoints were interpreted according to version 8.1. Data for S/R interpretations were exported from the ultra-rapid AST system. The system was only calibrated to distinguish S from I & R. Clinical AST results (S/I/R interpretations for disk diffusion on agar) for the clinical isolates used in the study were retrieved from the Laboratory Information System (LIS). No other patient information was obtained from the LIS. The results were collated alongside each other into an Excel software. Discrepant results obtained with the two AST methods were categorized as minor error (mE; ultra-rapid AST system = S or R and reference method = I); major error (ME; ultra-rapid AST system = R and reference method = S); very major error (VME; ultra-rapid AST system = S and reference method = R).

## RESULTS

The present study investigated the performance of the ultra-rapid AST system when assays were run for 30 min. Ultra-rapid AST was first performed directly on BC bottles. This involved prospectively testing BCs that signaled positive and were identified to contain *E. coli* (*n* = 59). The AST results were obtained in 30 min for 51/59 (86%) samples whereas 8/59 (14%) had invalid test result due to technical errors.

For the *E. coli*-positive BC bottles that obtained an AST result from the ultra-rapid AST system, the system provided concordant results with reference method in 47 out of 51 tests for Ciprofloxacin (Fig. 1A), and in 43 out of 51 tests for Gentamicin (Fig. 1B) within 30 minutes of starting the assay.

**Fig. 1.**
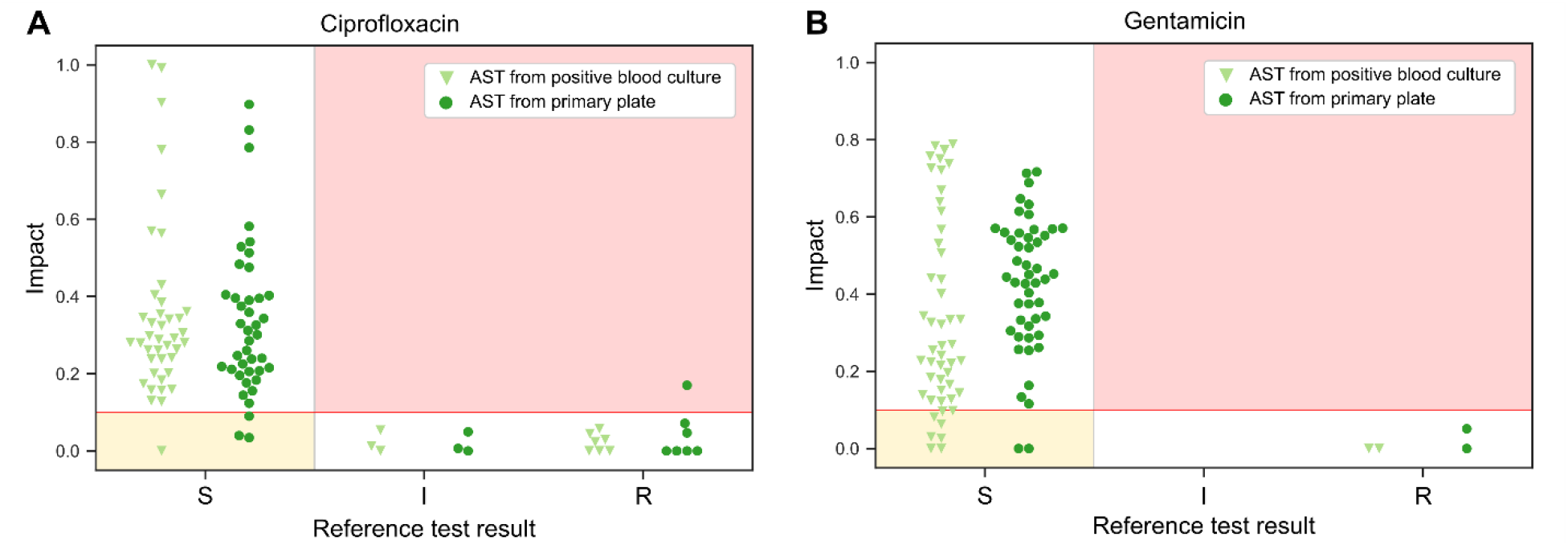
Measurement of *E. coli* growth rate impact in the presence of Ciprofloxacin and Gentamicin with the ultra-rapid AST system at 30 min assay time. The growth rate impact, defined as reduction of normalized growth rate at the decision time (30 min from assay start), is presented for tests performed on samples from positive blood cultures (triangles) and for tests performed on colonies from primary plates of isolates of the same samples (circles) in the presence of Ciprofloxacin (A) and Gentamicin (B). Growth rate impact is compared to the S/I/R AST results obtained by the reference method for the same samples. The red lines represent the susceptibility threshold of the test, the red regions in the plots represent false susceptible results, and the yellow regions represent false resistant results compared to the reference method.

In second part of the study where the ultra-rapid AST was performed on clinical *E. coli* isolates from agar plates, the ultra-rapid AST provided concordant results with reference method in 44 out of the 51 tests for Ciprofloxacin (Fig. 1A) and 49 out of 51 the tests for Gentamicin (Fig. 1B) within 30 minutes. When phenotypic responses for these tests were investigated over a time course up to 60 min, the number of results in concordance with the reference test increased to 47/51 for Ciprofloxacin (Fig. 2A) and to 51/51 (100%) for Gentamicin (Fig. 2B) with the longer assay time

**Fig. 2.**
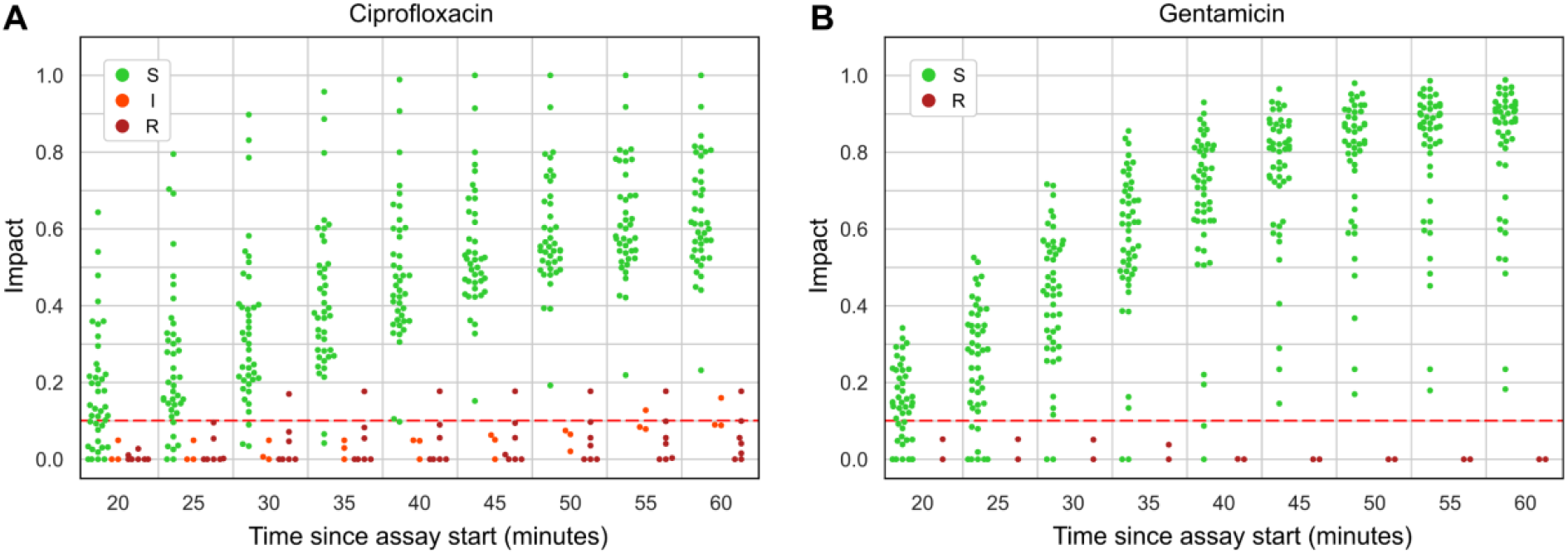
Measurement of *E. coli* growth rate impact in the presence of Ciprofloxacin and Gentamicin with the ultra-rapid AST system over time course. S/I/R classifications of E. coli isolates taken from agar plates based on their growth rate impact measured by the ultra-rapid AST system over 20-60 min in the presence of Ciprofloxacin (A) and Gentamicin (B). The growth rate impact, defined as reduction of normalized growth rate at the decision time, are dependent of the time from assay start. The red dashed line represents the susceptibility threshold for 30 minutes of assay time.

Discrepant results between the ultra-rapid method and disk diffusion are shown in Table 1.

**Table 1:** Discrepancies between AST Results obtained by ultra-rapid method and disk diffusion.

|  | Positive blood culture bottles |  |  |  |  | Agar Plate Isolates |  |  |  |  |  |  |  |  |  |
| --- | --- | --- | --- | --- | --- | --- | --- | --- | --- | --- | --- | --- | --- | --- | --- |
|  | (30min) |  |  |  |  | 30min |  |  |  |  | 60min |  |  |  |  |
| Abx | Total# | VME | ME | mE | %CA | Total# | VME | ME | mE | %CA | Total# | VME | ME | mE | %CA |
| CIP | 51 | 0 | 1 | 3 | 92 | 51 | 1 | 3 | 3 | 86 | 51 | 1 | 0 | 3 | 92 |
| GEN | 51 | 0 | 8 | 0 | 84 | 51 | 0 | 2 | 0 | 96 | 51 | 0 | 0 | 0 | 100 |
| Overall | 102 | 0 | 9 | 3 | 88 | 102 | 1 | 5 | 3 | 91 | 102 | 1 | 0 | 3 | 96 |

Abx: Antibiotic, VME: Very Major Error, ME: Major Error, mE: Minor Error, CA%: Categorical Agreement Percentage.

## DISCUSSION

The present study shows for the first time that the ultra-rapid AST can be performed directly from positive blood cultures in 30 min, with reliable accuracy for susceptibility testing of E. coli which is one of the leading causes of sepsis. The antibiotics tested were ciprofloxacin (92% CA) and gentamicin (84% CA) that are two distinct classes of antibiotics commonly used in the treatment of infections with E. coli. The ultra-rapid AST method showed similar high accuracy for Ciprofloxacin (86% CA) and Gentamicin (96% CA) when it is used on colonies from agar plates in 30 min. It is reasonable to suggest that the possibility to obtain information on AST in 30 min can be revolutionary in treatment of patients with BSI and other invasive infections. The turnaround time of 30 min enables implementation of the method in clinical routine not only for 24/7 use but also for laboratories with limited work hours. A 30 min method can provide actionable results in ∼94% of work hours in an 8am-to-5pm laboratory, while a 4 h tests can only generate actionable results in ∼55 % of the same work hours.

The present results obtained from positive blood cultures are in line with the previous study that was performed on uropathogenic *E. coli* isolates spiked into culture medium [9] despite major differences in sample matrix between the two studies. One of the key differences between these two studies is that present samples from blood culture bottles have a more complex sample matrix than the spiked samples used in the previous study, which did not contain all the blood components that are present in the blood culture bottles.

Longer assay duration was tested to investigate the impact of different decision times on the accuracy of the result. Analysis of different decision time points indicates that as the turnaround time increases up to 60 minutes, magnitude of impact differentiates resistant and susceptible population better. This, for the tests done from colonies on agar plates, increased the accuracy of AST results for Ciprofloxacin from 86% at 30 minutes, to 92% at 60 minutes. For Gentamicin, the increase in accuracy was from 96% at 30 minutes, to 100% at 60 minutes. Instead of simply interpreting the present data as a requirement for longer test durations, it could be considered that the data from the present ultra-rapid AST can be obtained earlier than 60 min by developing different thresholds of susceptibility for different time points. What is obvious is that there are clearly susceptible isolates detected as early as 20 minutes. Therefore, one can see that potentially a better utility of this test could be to have a time-dependent susceptibility cut-off and report clear susceptibility results as early as possible. A similar approach was taken with the EUCAST rapid AST method, which is a disk diffusion-based method adapted for AST directly on positive blood cultures, where the impact of test duration on AST accuracy is highlighted and multiple reading times are chosen with different thresholds to report clear results as early as possible while holding off the results falling into an uncertainty region, which gets smaller as the time goes from 4 hours to 8 hours [12].

The present ultra-rapid AST method can be improved by calibrating the data analysis algorithm using a large set of isolates to decide optimum cut-off thresholds for different time points and it can be used in ultra-rapid detection of susceptibility results between 20 to 60 minutes depending on susceptibility/resistance level of the bacteria tested.

Although the provision of a rapid means of identifying bacteria in the sample is not part of the ultra-rapid AST method. This is a strength instead of a weakness against utility of the test. Many laboratories already have implemented a multitude of different rapid identification methods, and by the addition of this method for ultra-rapid AST, they will obtain an actionable AST result in parallel. Even if one of these identification approaches fails or was not applicable to their sample, users can choose another rapid identification method without compromising AST outcome.

In conclusion, the present study shows that performing ultra-rapid phenotypic AST directly from positive blood culture bottles is possible and that the results can be obtained as early as in 30 min. Similarly, the method can be implemented on colonies from agar plates. Further studies implementing the ultra-rapid AST method on other species and antibiotics are warranted.

## Data Availability

The data presented in this study are openly available upon request.

## Statements and Declarations

### Ethics approval

Ethical review and approval were waived for this study, due to the use of leftover samples that were anonymized. The test results were not sent to the clinicians and the patient charts were not analyzed.

### Consent to participate

Written informed consent from the patient(s) were not necessary as described above.

### Consent for publication

All authors have read and agreed to the published version of the manuscript.

### Availability of data and material

The data presented in this study are openly available upon request.

### Code availability

Software to analyze is proprietary software of Astrego Diagnostics AB.

### Authors’ contributions

Conceptualization: Özden Baltekin; Methodology: Özden Baltekin; Formal analysis: Özden Baltekin, Erik Wistrand-Yuen; Investigation: Alexander T. A. Johnsson, Berivan Mert; Resources: Kajsa Nilsson, Lovisa Söderberg, Volkan Özenci; Data curation: Kajsa Nilsson, Lovisa Söderberg, Özden Baltekin; Writing – original draft: Özden Baltekin, Volkan Özenci; Writing –review & editing: Özden Baltekin, Volkan Özenci, Alicia Y. W. Wong; Supervision: Özden Baltekin, Volkan Özenci; Project administration: Özden Baltekin, Volkan Özenci.

## Acknowledgements

We thank the blood culture section at the Department of Clinical Microbiology at Karolinska University Hospital Huddinge, Stockholm, Sweden for their help and expertise with obtaining the clinical blood culture samples used for the present study.

This paper describes the use of an early prototype of Astrego’s AST System for the specific application. The conception of the Astrego’s technology, as well as development of the prototype AST System (analyzer, disposable cartridge, and analysis software) were made by several scientists and engineers that are not all authors of this paper.

## SUPPLEMENTARY MATERIAL

### Analysis of growth rate

The image analysis and growth rate calculations were performed similarly to as previously described (Baltekin et al. 2017). The instantaneous growth rates were calculated using a sliding window of 15 minutes, and for each antibiotic treatment the growth rate was normalized by the average growth rate of the reference treatment. To exclude debris and non-growing cells from the analysis, traps with an absolute growth rate of less than 0.005 divisions per minutes or a total change in length of less than 3 pixels during the assay were excluded. The normalized growth rate as a function of time was fitted to the sigmoid function y = L / (1 + exp(-k(t -t0))) + b using the “optimize” module of SciPy 1.4.1 in the Python programming language. The fit with the lowest sum of square residuals was chosen using the trust region reflective least square algorithm implemented in the “curve fit” function of the module. The growth rate impact was calculated as the reduction of normalized growth rate at the decision time based on the curve fitted to the data (Supplementary Figure.1)

**Supplementary Figure 1.**
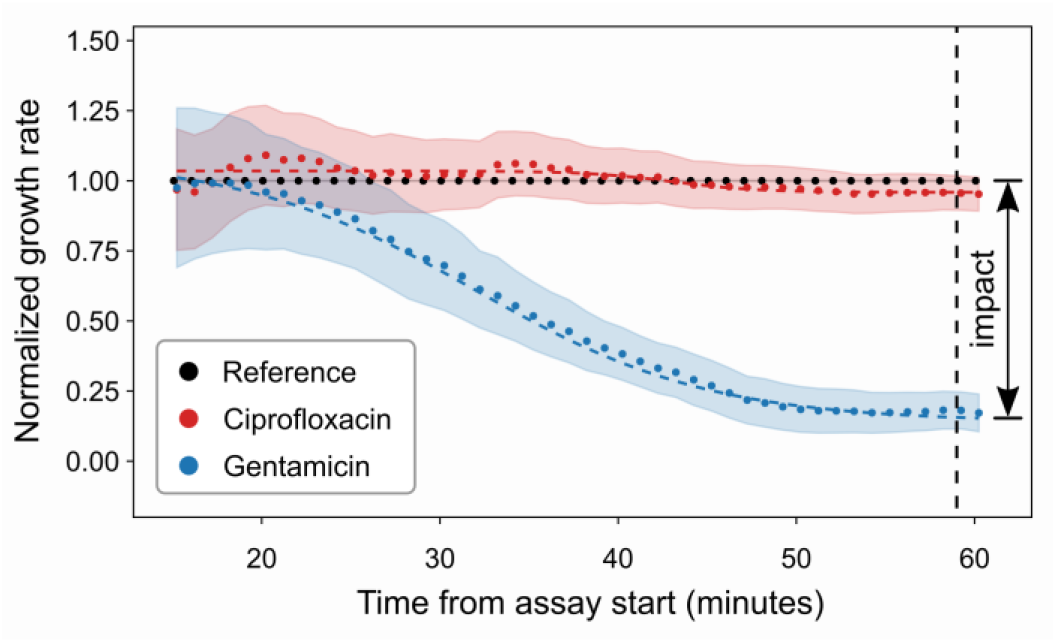
Definition of growth impact. Example assay results obtained from the ultra-rapid AST method. The normalized growth rates of an *E. coli* isolate in the presence of Ciprofloxacin (red) and Gentamicin (blue) are shown as filled circles over time. Sigmoid fitted curves are shown as dashed lines. The shaded regions show the 99.9% confidence intervals of the mean. The growth impact for each drug is defined as the reduction of the normalized growth rate at the decision time based on the fitted curve. The arrow on the right in the figure indicates the impact on bacteria growth for Gentamicin at 60 minutes.

**Supp. Table 1:** Control strains from Astrego Diagnostics AB.

| *E. coli* strain | Ciprofloxacin (MIC mg/L) | Gentamicin (MIC mg/L) |
| --- | --- | --- |
| ADS-121 | R (16) | S (<2) |
| ADS-268 | S (<0.5) | S (<2) |
| ADS-566 | S (<0.5) | S (<2) |
| ADS-434 | S (<0.5) | S (<2) |

S, susceptible; R, resistant

